# Head-to-head comparison of SARS-CoV-2 antigen-detecting rapid test with self-collected anterior nasal swab versus professional-collected nasopharyngeal swab

**DOI:** 10.1101/2020.10.26.20219600

**Authors:** Andreas K. Lindner, Olga Nikolai, Franka Kausch, Mia Wintel, Franziska Hommes, Maximilian Gertler, Lisa J. Krüger, Mary Gaeddert, Frank Tobian, Federica Lainati, Lisa Köppel, Joachim Seybold, Victor M. Corman, Christian Drosten, Jörg Hofmann, Jilian A. Sacks, Frank P. Mockenhaupt, Claudia M. Denkinger

## Abstract

**Background:** Two antigen-detecting rapid diagnostic tests (Ag-RDTs) are now approved through the WHO Emergency Use Listing procedure and can be performed at the point-of-care. However, both tests use nasopharyngeal (NP) swab samples. NP swab samples must be collected by trained healthcare personnel with protective equipment and are frequently perceived as uncomfortable by patients.

**Methods:** This was a manufacturer-independent, prospective diagnostic accuracy study with comparison of a supervised, self-collected anterior nose (AN) swab sample with a professional-collected NP swab sample, using a WHO-listed SARS-CoV-2 Ag-RDT, STANDARD Q COVID-19 Ag Test (SD Biosensor), which is also being distributed by Roche. The reference standard was RT-PCR from an oro-/nasopharyngeal swab sample. Percent positive and negative agreement as well as sensitivity and specificity were calculated.

**Results:** Among the 289 participants, 39 (13.5%) tested positive for SARS-CoV-2 by RT-PCR. The positive percent agreement of the two different sampling techniques for the Ag-RDT was 90.6% (CI 75.8-96.8). The negative percent agreement was 99.2% (CI 97.2-99.8). The Ag-RDT with AN sampling showed a sensitivity of 74.4% (29/39 PCR positives detected; CI 58.9-85.4) and specificity of 99.2% (CI 97.1-99.8) compared to RT-PCR. The sensitivity with NP sampling was 79.5% (31/39 PCR positives detected; CI 64.5-89.2) and specificity was 99.6% (CI 97.8-100). In patients with high viral load (>7.0 log _10_ RNA SARS-CoV2/swab), the sensitivity of the Ag-RDT with AN sampling was 96% and 100% with NP sampling.

**Conclusion:** Supervised self-sampling from the anterior nose is a reliable alternative to professional nasopharyngeal sampling using a WHO-listed SARS-CoV-2 Ag-RDT. Considering the ease-of-use of Ag-RDTs, self-sampling and potentially patient self-testing at home may be a future use case.

## To the Editor

A number of antigen-detecting rapid diagnostic tests (Ag-RDTs) for SARS-CoV-2 are now commercially available and can result in rapid decisions on patient care, isolation and contact tracing at the point-of-care [1]. Two Ag-RDTs using nasopharyngeal (NP) swab samples meet WHO targets and are now approved through the WHO Emergency Use Listing procedure [2-4].

NP swab samples are frequently perceived as uncomfortable by patients and must be collected by trained healthcare personnel with protective equipment. Evidence supports the use of alternative sampling methods for RT-PCR, including anterior nasal (AN) swabs collected by patients and some tests have received regulatory approval with AN samples [5, 6]. Considering the ease-of-use of Ag-RDTs, a reliable simple sampling method would not only allow self-sampling, but may also pave the way for self-testing.

The primary objective of this prospective diagnostic accuracy study was a head-to-head comparison (positive and negative percent agreement) of a supervised, self-collected AN swab sample with a health care worker (professional)-collected NP swab sample, using a WHO-listed SARS-CoV-2 Ag-RDT against the reference standard RT-PCR collected from a NP/oropharyngeal (OP) swab. The secondary objective was to assess sensitivity and specificity for different sampling techniques with Ag-RDT. The study was continued until at least 30 positive NP swab samples according to Ag-RDT were obtained. This manufacturer-independent study was conducted in partnership with the Foundation of Innovative New Diagnostics (FIND), the WHO collaborating centre for COVID-19 diagnostics.

The study protocol was approved by the ethical review committee at Heidelberg University Hospital for the study site in Berlin (registration number S-180/2020). The study took place at the ambulatory SARS-CoV-2 testing facility of Charité University Hospital (Charité-Universitätsmedizin Berlin, Germany) from 23 September to 14 October 2020. The study enrolled adults at high risk for SARS-CoV-2 infection according to clinical suspicion. Participants were excluded if either of the swabs for the Ag-RDT or the RT-PCR reference standard could not be collected.

Participants underwent first an instructed, self-collected bilateral AN swab for the Ag-RDT. Verbal instruction was given to insert the swab horizontally 2-3 cm into the nostril and rotate it for 15 seconds against the nasal walls on each side. Deviations from the instructed technique were recorded. Subsequently, a combined OP/NP swab (eSwab from Copan) as per institutional recommendations for RT-PCR, and a separate NP swab for the Ag-RDT were taken from different sides of the nose. The samples for the Ag-RDTs were collected using the swab provided by the manufacturer within the test kit.

The Ag-RDT evaluated in this study was the STANDARD Q COVID-19 Ag Test (SD Biosensor, Inc. Gyeonggi-do, Korea; henceforth called STANDARD Q) [7], which is also being distributed by Roche [8]. The test uses the lateral flow assay principle in a cassette-based format with a visual read-out after 15-30 minutes. The manufacturer’s instructions for use were followed. The Ag-RDTs were performed directly after sampling (within 60 minutes) at point-of-care by study physicians. The Ag-RDT results were interpreted by two operators, each blinded to the result of the other. The second reader was also blinded to the second Ag-RDT results of individual patients. The visual read out of the Ag-RDT test band was categorized on a semi-quantitative scale as negative, weak positive, positive and strong positive.

The Roche Cobas SARS-CoV-2 assay (Pleasanton, CA United States) or the SARS-CoV-2 E-gene assay from TibMolbiol (Berlin, Germany) were performed for RT-PCR according to routine procedures at the central laboratory. Viral RNA concentrations were calculated using assay specific CT-values, based on external calibrations curves [9, 10]. Staff performing the Ag-RDTs were blinded to results of RT-PCR tests and vice versa.

Of 303 patients invited, 289 (95.4%) consented to participate. Two patients were excluded as both swabs for the Ag-RDT could not be obtained. The average age of participants was 34.7 years (Standard Deviation [SD] 11.0) with 42.9% female and 19.0% having comorbidities. On the day of testing, 97.6% of participants had one or more symptoms consistent with COVID-19. Duration of symptoms at the time of presentation on average was 4.4 days (SD 2.7). Among the 289 participants, 39 (13.5%) tested positive for SARS-CoV-2 by RT-PCR (Table 1).

**TABLE 1.** Antigen-detecting RDT results with a supervised self-collected anterior nasal (AN) swab and with a professional-collected nasopharyngeal (NP) swab in RT-PCR positive patients from combined oro-/nasopharyngeal swab. CT-values and viral load (in descending order) of the paired RT-PCR samples are shown, as well as the duration of symptoms per patient. The positive percent agreement between AN and NP samples on Ag-RDT, as well as the respective sensitivities compared to RT-PCR are shown.

| No. | AN swab<br>self-collected<br>SD Q Ag-RDT | NP swab<br>prof.-collected<br>SD Q Ag-RDT | OP/NP swab<br>RT-PCR |  | Symptom<br>duration<br>(days) |
| --- | --- | --- | --- | --- | --- |
|  |  |  | CT value | Viral load<sup>3</sup> |  |
| 1 | pos (+++) | pos (+++) | 17.33<sup>1</sup> | 9.92 | 2 |
| 2 | pos (++) | pos (+++) | 17.86<sup>1</sup> | 9.76 | 1 |
| 3 | pos (+++) | pos (+++) | 18.01<sup>1</sup> | 9.71 | 1 |
| 4 | pos (++) | pos (+++) | 18.31<sup>1</sup> | 9.62 | 3 |
| 5 | pos (+++) | pos (+++) | 18.40<sup>1</sup> | 9.60 | 3 |
| 6 | pos (+++) | pos (+++) | 18.76<sup>1</sup> | 9.49 | 4 |
| 7 | pos (+++) | pos (+++) | 18.77<sup>1</sup> | 9.49 | 5 |
| 8 | pos (+++) | pos (+++) | 18.78<sup>1</sup> | 9.49 | 5 |
| 9 | pos (+++) | pos (+++) | 19.05<sup>1</sup> | 9.41 | 3 |
| 10 | pos. (+++) | pos. (+++) | 19.40<sup>1</sup> | 9.30 | 2 |
| 11 | neg. | pos (+++) | 19.66<sup>1</sup> | 9.23 | 1 |
| 12 | pos (+++) | pos (+++) | 20.32<sup>1</sup> | 9.03 | 3 |
| 13 | pos (++) | pos (+++) | 20.44<sup>1</sup> | 9.00 | 2 |
| 14 | pos (++) | pos (++) | 20.54<sup>1</sup> | 8.96 | 5 |
| 15 | pos (+++) | pos (+++) | 17.81<sup>2</sup> | 8.86 | 4 |
| 16 | pos (+++) | pos (+++) | 21.09<sup>1</sup> | 8.80 | 4 |
| 17 | pos (+++) | pos (+) | 18.62<sup>2</sup> | 8.62 | 4 |
| 18 | pos (+) | pos (++) | 21.87<sup>1</sup> | 8.57 | 7 |
| 19 | pos (++) | pos (+++) | 22.05<sup>1</sup> | 8.52 | 2 |
| 20 | pos (++) | pos (+++) | 19.34<sup>2</sup> | 8.41 | 5 |
| 21 | pos (+++) | pos (+++) | 19.47<sup>2</sup> | 8.37 | 6 |
| 22 | pos (+++) | pos (+++) | 22.60<sup>1</sup> | 8.36 | 6 |
| 23 | pos (+++) | pos (++) | 23.66<sup>1</sup> | 8.04 | 6 |
| 24 | pos (+) | pos (++) | 26.42<sup>1</sup> | 7.23 | 5 |
| 25 | pos (+++) | pos (+++) | 26.77<sup>1</sup> | 7.12 | 5 |
| 26 | neg. | neg. | 24.25<sup>2</sup> | 6.96 | 10 |
| 27 | pos (++) | pos (+++) | 24.77<sup>2</sup> | 6.80 | 4 |
| 28 | pos (+++) | pos (++) | 25.29<sup>2</sup> | 6.64 | 2 |
| 29 | pos (+) | pos (++) | 29.33<sup>1</sup> | 6.36 | 5 |
| 30 | neg. | neg. | 29.56<sup>1</sup> | 6.30 | 3 |
| 31 | neg. | neg. | 29.95<sup>1</sup> | 6.18 | 3 |
| 32 | pos (+) | pos (+) | 30.25<sup>1</sup> | 6.09 | 4 |
| 33 | neg. | neg. | 27.81<sup>2</sup> | 5.90 | 8 |
| 34 | pos (++) | pos (+) | 31.20<sup>1</sup> | 5.81 | 8 |
| 35 | neg. | pos (+) | 31.61<sup>1</sup> | 5.69 | 10 |
| 36 | neg. | neg. | 32.58<sup>1</sup> | 5.40 | 10 |
| 37 | neg. | neg. | 32.86<sup>1</sup> | 5.32 | 2 |
| 38 | neg. | neg. | 34.62<sup>1</sup> | 4.80 | 7 |
| 39 | neg. | neg. | 35.53<sup>1</sup> | 4.53 | 14 |
|  | Sensitivity<br>29/39 (74.4%) | Sensitivity<br>31/39 (79.5%) | <sup>1</sup> Roche Cobas SARS-CoV-2 assay (E-gene, T2 target)<br><sup>2</sup> TibMolbiol assay, E-gene target.<br><sup>3</sup> log<sub>10</sub> RNA SARS-CoV2/swab<br><sup>4</sup> including 2 false positives on AN and 1 on NP |  |  |
|  | Positive percent agreement<sup>4</sup><br>90.6% (CI 75.8-96.8) |  |  |  |  |

**Abbreviations:** No., patient number; SD Q, STANDARD Q COVID-19 Ag Test (SD Biosensor); Ag-RDT, antigen-detecting rapid diagnostic test; AN, anterior nasal; NP, nasopharyngeal; OP, oropharyngeal; CT, cycle threshold; RT-PCR, reverse transcription-polymerase chain reaction; neg., negative; pos (+), weak positive; pos. (++), positive; pos. (+++), strong positive.

No invalid tests were observed on either AN or NP samples. Two patients were detected by NP but not by AN sampling. No patient was detected by AN sampling only. The positive percent agreement was 90.6% (CI 75.8-96.8; including 2 false positive results with AN and 1 with NP). The negative percent agreement was 99.2% (CI 97.2-99.8). Inter-rater reliability was near perfect with kappa of 0.98 for AN and 1.0 for NP samples. The semi-quantitative read-out was more often higher for the NP samples (9 higher on NP, 4 higher on AN). Of the two patients detected by NP but not by AN sampling, one patient collected the AN swab only with gentle rotation, the second presented 10 days post symptom onset with a low viral load (Table 1).

The STANDARD Q Ag-RDT with AN sampling showed a sensitivity of 74.4% (29/39 PCR positives detected; CI 58.9-85.4) and specificity of 99.2% (CI 97.1-99.8) compared to RT-PCR. The sensitivity with NP sampling was 79.5% (31/39 PCR positives detected; CI 64.5-89.2) and specificity was 99.6% (CI 97.8-100). In patients with high viral load (>7.0 log_10_ RNA SARS-CoV2/swab), the sensitivity of the Ag-RDT with AN sampling was 96% (24/25 PCR positives detected; CI 80.5-99.8) and 100% (25/25 PCR positives detected; CI 86.7-100) with NP sampling. In contrast, the Ag-RDT frequently did not detect patients with lower viral load or with symptoms >7 days (Table 1). For most patients, the application of the flexible swab (meant for NP swab collection) in the anterior nose appeared unpleasant due to a tickling sensation and led to frequent sneezing.

The strengths of the study are the rigorous methods, including standardized sampling, two independent blinded readers and an additional semi-quantitative assessment of Ag-RDT results. The cohort was representative, judging from the comparable sensitivity observed in the recent independent validation study of STANDARD Q (sensitivity 76.6%; CI 62.8-86.4) [4]. The study is limited as it was performed in a single centre. Also, the NP swab was usually rotated against the nasopharyngeal wall for less time than recommended by the manufacturer, which may have a negative impact on the sensitivity of the Ag-RDT with NP sampling, but also reflects the difficulty of collection of this sample type.

In conclusion, this study demonstrates that supervised self-sampling from the anterior nose is a reliable alternative to professional nasopharyngeal sampling with STANDARD Q. The data will contribute to WHO recommendations for use of this test. Considering the ease-of-use of Ag-RDTs, self-sampling and potentially patient self-testing at home may be a future use case. If such testing could be repeated frequently and immediately ahead of situations when transmissions are likely to occur, self-testing with Ag-RDTs may have a significant impact on the pandemic. Further implementation studies on optimized self-sampling techniques and swabs (e.g. less flexible sponge swab) and the correct performance/interpretation of the test by patients themselves, are urgently needed to drive self-testing to scale.

## Data Availability

All raw data and analysis code are available upon a request to the corresponding author.

## Acknowledgements

Heike Rössig, Chiara Manon Rohardt, Claudia Hülso, Elisabeth Linzbach, Susen Burock, Katja von dem Busche, Stephanie Patberg, Melanie Bothmann, Zümrüt Tuncer, Stefanie Lunow, Beate Zimmer, Astrid Barrera Pesek, Sabrina Pein, Nicole Buchholz, Verena Haack, Oliver Deckwart.

## Author contributions

AKL, LJK, FL and CMD designed the study and developed standard operating procedures. AKL and ON implemented the study design, enrolled patients, performed laboratory work and led the writing of the manuscript. FPM and JS coordinated and supervised the study site. FK, MW, FH enrolled patients. MGe coordinated the testing facility. MGa, LK and FT led the data analysis. VC, JH and CD were responsible for PCR testing and contributed to the interpretation of the data. JAS supported the study design setup and the interpretation of the data. CMD was the principle investigator of the study. All authors have reviewed the manuscript.

## Data availability

All raw data and analysis code are available upon a request to the corresponding author.

